# Prone Cardiopulmonary Resuscitation: A Rapid Scoping and Expanded Grey Literature Review for the covid-19 Pandemic

**DOI:** 10.1101/2020.05.21.20109710

**Authors:** Matthew J Douma, Ella Mackenzie, Tess Loch, Maria C. Tan, Dustin Anderson, Christopher Picard, Lazar Milovanovic, Domhnall O’Dochartaigh, Peter G Brindley

## Abstract

**Prone Cardiopulmonary Resuscitation:** A Rapid Scoping and Expanded Grey Literature Review for the COVID-19 Pandemic

**Aim:** To rapidly identify and summarize the available science on prone resuscitation. To determine the value of undertaking a systematic review on this topic; and to identify knowledge gaps to aid future research, education and guidelines.

**Methods:** This review was guided by specific methodological framework and reporting items (PRISMA-ScR). We included studies, cases and grey literature regarding prone position and CPR/cardiac arrest. The databases searched were MEDLINE, Embase, CINAHL, Cochrane CENTRAL, Cochrane Database of Systematic Reviews, Scopus and Google Scholar. Expanded grey literature searching included internet search engine, targeted websites and social media.

**Results:** Of 453 identified studies, 24 (5%) studies met our inclusion criteria. There were four prone resuscitation-relevant studies examining: blood and tidal volumes generated by prone compressions; prone compression quality metrics on a manikin; and chest computed tomography scans for compression landmarking. Twenty case reports/series described the resuscitation of 25 prone patients. Prone compression quality was assessed by invasive blood pressure monitoring, exhaled carbon dioxide and pulse palpation. Recommended compression location was zero-to-two vertebral segments below the scapulae. Twenty of 25 cases (80%) survived prone resuscitation, although few cases reported long term outcome. Seven cases described full neurological recovery.

**Conclusion:** This scoping review did not identify sufficient evidence to justify a systematic review or modified resuscitation guidelines. It remains reasonable to initiate resuscitation in the prone position if turning the patient supine would lead to delays or risk to providers or patients. Prone resuscitation quality can be judged using end-tidal CO2, and arterial pressure tracing, with patients turned supine if insufficient.

## Introduction

For patients to have the best chance of surviving sudden cardiac arrest, high quality chest compressions and defibrillation need to be initiated without delay.[1] Traditionally, cardiopulmonary resuscitation (CPR) is taught and performed in the supine position, but even short delays-such as would occur if proned patients are first turned to the supine position - can be clinically detrimental.[2] The question of whether to supine patients before initiating resuscitation has long been relevant for out-of-hospital arrest patients found face down, and for intraoperative arrests during prone surgeries i.e. spine surgery, neurosurgery, retroperitoneal surgery. More recently, there has been an increase in prone mechanical ventilation in Intensive Care Unit (ICU) patients, since the 2013 Prone Positioning in Severe Acute Respiratory Distress Syndrome trial.[3] Since the covid-19 pandemic, however, the issue of prone resuscitation has become especially important.[4] This is because, while we await robust evidence, prone positioning has been recommended in both confirmed and suspected cases, and in severely-ill/sedated patients and less ill/awake patients.[5–9]

‘Flipping’ a patient from prone to supine, especially during an emergency, comes with potential risks to both patient and staff.[10] Other than time delay, there could be endotracheal tube dislodgement, disconnection of vascular lines, disconnection of ventilation tubing, and staff contamination. There is also the mechanical challenge of turning obese patients, and potential injury to staff and patients. Especially relevant during pandemics, rolling a patient means that extra people (often six in total) enter a potentially contaminated space. There means extra delays until all of these staff have appropriately donned protective equipment, and the cumulative risks associated with doffing.

While almost all practitioners will supine patients before attempting an advanced airway, there is an initial period where compressions and defibrillation are performed before airway capture or supplementary breaths. Moreover, resuscitation guidelines have acknowledged that there are incidences where initiation of non-supine resuscitation is appropriate.[11] To date, most frontline clinicians have limited knowledge of the literature surrounding prone resuscitation. This is noteworthy now that proning is increasing. The goal of this scoping review is to assist clinicians forced to decide, in the midst of an emergency, whether CPR should be initiated with the patient ‘proned’, or whether the risks and delays of ‘supine-ing’ are unavoidable. More specifically, our objectives are: i) to examine the extent of research into prone resuscitation, ii) determine the value of undertaking a full systematic review, iii) summarize and disseminate research related to prone resuscitation, iv) and identify strategic gaps in the literature to aid future research, education and guidelines.

## Methods

This scoping review followed the methodological framework of Arksey and O’Malley and the enhancements of Levac et al.[12,13] It incorporated six steps: i) research question identification, ii) relevant study identification, iii) selection of studies, iv) charting the data, v) collate, summarize and report results, and vi) contributor provided references. Furthermore, this review adheres to the Preferred Reporting Items for Systematic Reviews and Meta-Analyses extension for scoping reviews (PRISMA-ScR).[14] Our findings are presented in tabular format, accompanied by narrative summaries.

### Search strategy and inclusion criteria

The search strategy aimed to locate published and unpublished primary studies, reviews and grey literature such as policy, procedure and social media. We constructed a minimally-restrictive review question following the “PICO” format: for (p) patients (or manikins or cadavers) in the prone position in at the time of cardiac arrest (real or simulated), (i) who receive cardiopulmonary resuscitation, (c) no comparison criteria, and (o) any outcome. Full strategy is available in Appendix 1. Studies were eligible if they were peer-reviewed and examined or reported any characteristic or outcome of interest to prone CPR. Both qualitative and quantitative studies were eligible for inclusion. Studies using animal models of prone CPR were excluded.

### Data sources

A systematic literature search was conducted by a health sciences librarian (MCT) using the following bibliographic databases: Ovid Medline ALL, Ovid Embase, EBSCOHost CINAHL Plus With full Text, Wiley Cochrane Central Register of Controlled Trials, Wiley Cochrane Database of Systematic Reviews, and Scopus. The search included a combination of controlled vocabulary (e.g., MeSH) and keywords representing the concepts of prone positioning and CPR/compressions. There were no limits on language, date, or study design. The MEDLINE search strategy was developed by the librarian and principal investigator (MJD), then translated for each database, and conducted on April 25, 2020. We also hand-searched bibliographies from previous systematic reviews for eligible studies. Multiple structured searches of Google Scholar were also performed and experts reviewed our search strategy and helped identify any missing articles.

Our systematic grey literature search strategy was designed by author MD and followed previously accepted, peer-reviewed, methods.[15] It consisted of four parts: i) internet search engine (Chrome anonymous browser for de-personalized Google search without geographical bias) (EM), ii) targeted website searching of emergency department, critical care and resuscitation organizations (TC), iii) grey literature database searching (KS & EM), and iv) social media platform searching, including blogs (DOD). Grey literature sources were identified using accepted resources.[16] The purpose of our grey literature search was: i) to identify relevant cases of prone resuscitation and inquire as to their publication and ii) gain insight from relevant policy, procedure and clinical governance documents. Our grey literature search strategy is detailed in Appendix 2.

### Study selection

References were imported to Covidence [Vertias Health Innovation, Melbourne, Australia] for deduplication, title, abstract, and full text screening. Authors DA, MJD, DOD and LM independently screened titles, abstracts and full text articles against the selection criteria. Disagreements were resolved by discussion and consensus.

### Data extraction and charting

Data were extracted by a single author (EM) and ratified by co-authors (TC and CP). Data were extracted into a Google Spreadsheet [Google, Mountain View California, USA]. An a priori data extraction table used to describe the characteristics of each study including: the authors(s), year of publication, study design, country, population, intervention and comparator (if applicable), major finding and outcome(s) examined. Data extracted from case reports/series included author, year, age, sex, diagnosis, type of surgery, initial rhythm, CPR quality and outcome (MJD and PGB).

## Results

The search process retrieved 327 references from bibliographic databases. Once duplicates were removed, 202 references remained. An additional 126 references were identified from other sources as per the methods described above. A further 59 duplicates were identified and removed, resulting in a total of 269 items for title and abstract screening. After applying the inclusion/exclusion criteria to titles/abstracts, 46 articles underwent full text screening. Subsequently, 22 more articles were excluded for the following reasons: the patient was returned to the supine position prior to initiating resuscitation (n=6), cardiac arrest could not be confirmed (n=6), no new data was presented (n=4), data was from a pilot of a study already included (n = 2), no resuscitation was attempted, or we were unable to obtain a full text article (n = 2). See PRISMA flow sheet (Figure 1) for more details. Overall, of 453 identified studies, 24 (5%) studies met our inclusion criteria and underwent extraction and narrative summary.[17-40]

**Figure 1.**
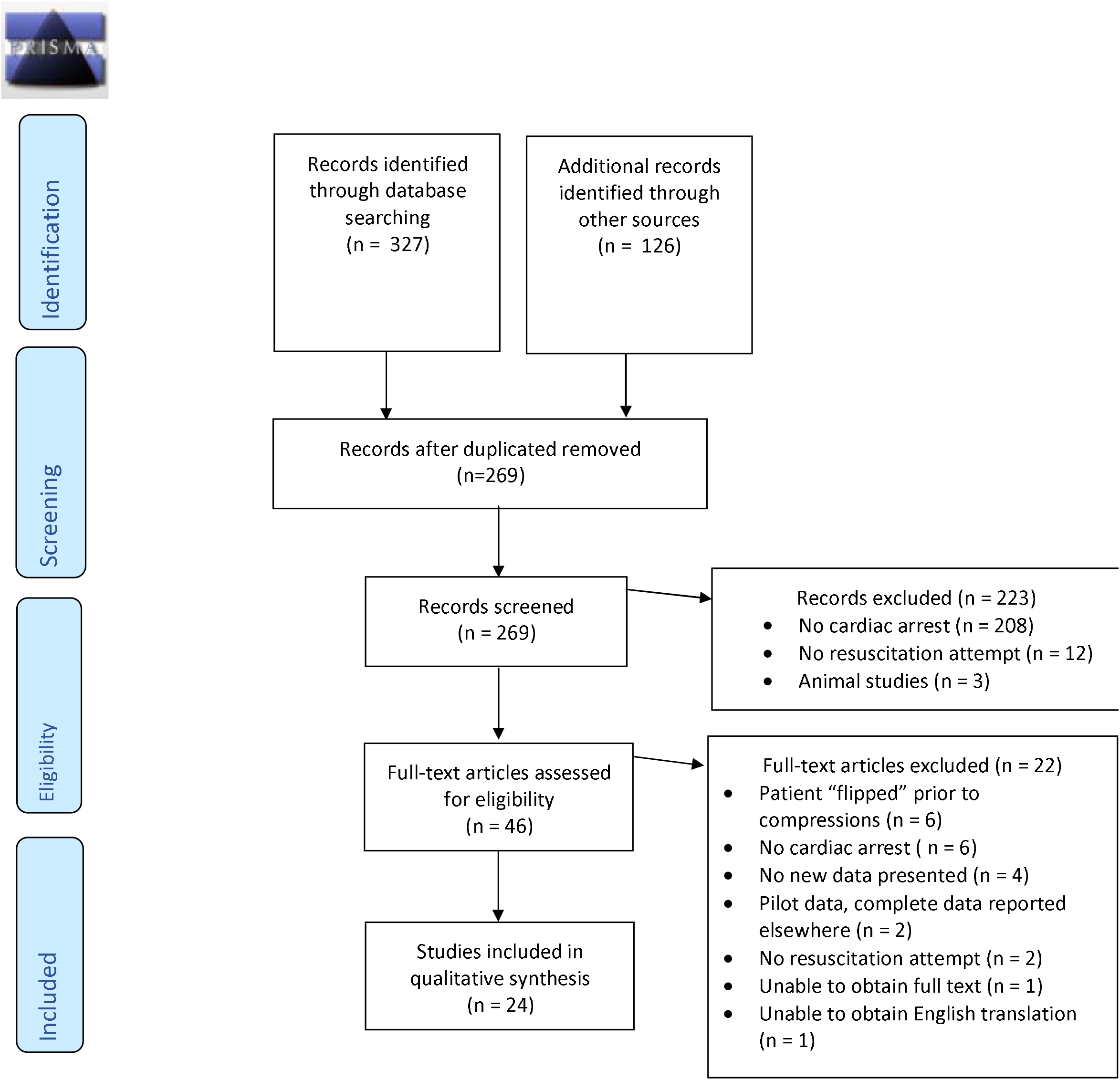
Prone Cardiopulmonary Resuscitation PRISMA Flow Diagram

### Study characteristics

Four original research studies met inclusion criteria: i) a feasibility study of blood pressure during supine versus prone compressions in six intensive care patients after prolonged cardiac arrest;[37] ii) a study of cadavers (n = 11) and healthy volunteers (n = 10) on blood pressures and tidal volumes generated by prone compressions;[38] iii) a prone compression quality manikin study;[39] and iv) a retrospective descriptive study of chest computed tomography scans of 100 patients to identify ideal prone compression hand position.[40] Refer to Table one for summary table of included research.

**Table one.** Details of included studies.

| **Table one. Details of included studies:** |  |  |  |  |
| --- | --- | --- | --- | --- |
| **Author, year** | **Design, Country** | **Population** | **Intervention / Comparator** | **Outcomes** |
| Atkinson, 2000 | Feasibility trial; Edinburgh, Scotland | 36 Registered Nurses | Nurses performed 100 compressions on a manikin with a gel-filled pad between sternum and standard examination couch. Landmark was midline ~2/3 up the body of the manikin i.e. "between imaginary scapula". | 3376 chest compressions: 1168 (34.6%) effective (4-5 cm compression), 1370 (40.6%) partially effective (2-4 cm), 838 (24.6%) ineffective (<2 cm depth). 22/36 (61%) nurses able to deliver effective compressions (mean of 53% compressions were effective). Wide variations in compression effectiveness. |
| Kwon, 2017 | Retrospective descriptive study; Seoul, Republic of Korea | 100 prone patients (54 male; 46 female; mean age 63 ± 12 years). | Single investigator, retrospective chest CT images of 100 prone patients: to identify the surface anatomical landmark, and approximate hand position during prone CPR. | Largest LV cross-sectional area is 0-2 vertebral segments below inferior angle of the scapulae in 86% of prone patients. |
| Mazer, 2003 | Pilot feasibility study, participants not blinded to blood pressure; New York, USA | 6 adult ICU patients enrolled over 1 year from CICU (n=4) and MICU (n=2); 4 male; 2 female. | 15min of supine compressions then rolled for 15min of prone compression (hands over the T7-10 vertebrae). | SBP improved 48 mmHg to 72 mmHg, MAP from 32mmHg to 46mmHg from supine to prone. No survivors and no ROSC. |
| Wei, 2006 | Part 1: cadavers. Part 2: healthy volunteers. Taiwan | Part 1: 11 cadavers (mean body weight 65.4 ± 9.0 kg). Part 2: 10 healthy volunteers (mean body weight 63.3 ± 14.7 kg) | Part 1: standard pre-cordial cardiac massage, then prone compressions. Part 2: respiratory assessment during prone compressions. | Part 1: Prone CPR resulted in SBP 79.4 ± 20.3 mmHg, and DBP 16.7 ± 10.3 mmHg. Supine CPR produced increase in SBP of 55.4 ± 20.3/13.0 ± 6.7 mmHg to (79 ± 20/17 ± 10) (P=0.028). Part 2: External back compression on volunteers generated a mean tidal volume of 399 ± 110 mL. |

Abbreviations: BP- Blood pressure; CICU- Cardiac intensive care unit; CPR- Cardiopulmonary resuscitation; CT- Computed tomography; DBP- Diastolic blood pressure; ETCO<sub>2</sub>- End tidal carbon dioxide; ETT- Endotracheal tube; ICU- Intensive care unit; IV- Intravenous ; LV- left ventricle ; MAP- Mean arterial pressure ; MICU- Medical intensive care unit ; PEA- Pulseless electrical activity; ROSC- Return of spontaneous circulation; TTE- Transthoracic echocardiography; SBP- Systolic blood pressure; SVT- Supraventricular tachycardia; VF- Ventricular fibrillation

### Case reports/series characteristics

Twenty case reports/series describe 25 prone patients undergoing resuscitation including compressions.[17-36] All but five occurred in an operating theatre.[22,36] Over half involved pediatric patients (n=14). [19,24,26–28,30,32,33,35,36] The arrest precipitant was incompletely described, but, five patients had air emboli,[26,28,29,33] three had cardiac decompensation,[17,25,35] three had airway obstruction,[32,36] two had excessive parasympathetic stimulation[27,32] and one had cardiac tamponade.[31] The most common initial rhythm was pulseless electrical activity (n = 11, 44%),[17–21,23–26,32] followed by asystole (n=4, 16%).[22,29,33] Refer to Table two for summary table of included cases.

**Table two.** Characteristics of Included Case Studies.

| **Author, year** | **Age** | **Sex** | **Diagnosis** | **Type of Surgery** | **Rhythm** | **Confirmation of CPR quality** | **Cause of arrest** | **Outcome (neurological ideally)** |
| --- | --- | --- | --- | --- | --- | --- | --- | --- |
| AlHarbi, 2020 | 80 | Male | Spine surgery | Posterior spinal fusion with laminectomy | PEA, asystole | ETCO<sub>2</sub> 10 mmHg with prone CPR but no palpable pulse. When switched to supine, ETCO<sub>2</sub> increased & had palpable CPR pulses | Intraoperative NSTEMI due to hypovolemia | Survived |
| Brock-Utne, 2011 | 28 | Male | Pelvic fractures | Open reduction internal fixation | PEA | Systolic pressures over 80 mmHg (from arterial wave form) | Pulmonary embolus | Survived (discharged from hospital with intact neurologic status) |
| Burki, 2017 | 6 | Female | Ventricle tumor excision | Posterior cranial fossa | PEA | Not discussed | Hemorrhagic shock | ROSC, died 5 days post-surgery |
| Chauhan, 2016 | 49 | Male | Prolapsed intervertebral disc | Lumbar discectomy | PEA, asystole | Not discussed | Unknown | Survived |
| Cho, 2010 | 18 | Female | Congenital posterior scoliosis | Deformity correction and fusion with instrumentation | PEA, VF | Not discussed | Unknown | Survived |
| Dequin, 1996 | 48 | Male | Pneumonia | Prone ICU ventilation | Asystole | Arterial BP >80/35mmHg throughout CPR | Unknown | Survived |
| Dooney, 2010 | 43 | Male | Left lower leg pain and weakness | L4-L5 microscopic discectomy | Bradycardic PEA to asystole | ETCO<sub>2</sub> =28mmHg | Unknown | Survived |
| Gueugniaud, 1995 | 15 | Male | Marfan's syndrome | Correction of a right dorsal and left lumbar scoliosis | PEA to asystole. Prone CPR to Supine CPR then ROSC | No palpable carotid pulse, ETCO<sub>2</sub><10mmHg | Unknown | Survived, extubated after 18hrs, full neurologic recovery |
| Haffner, 2010 | 81 | Male | Cerebellar hemorrhage | Craniotomy with acute cerebellar hemorrhage | PEA | ETCO<sub>2</sub> 33mmHg with compressions. Visible arterial BP waveform correlated with CPR | TTE confirmed intraoperative right cardiac decompensation | Vasopressors weaned day 2, recovered some neurologic function. Rearrested & died 1-month post discharge |
| Kalaria, 2017 | 1 | Female | Split cord malformation type1 with tethered cord | Tethered cord release and bone spur removal | Bradycardic PEA | Not discussed | Massive venous air embolus | Survived |
| Kaur, 2016 | 14 | Male | Koch's spine | Thoracic spine deformity correction | Severe bradycardia to asystole | Not discussed | Parasympathetic stimulation | CPRx4min then ROSC. Re-Arrested at 12hrs and died |
| Kelleher, 1995 | 6 months | Female | Achondroplasia, congenital cervical cord compression | Foramen magnum decompression | Multifocal PVCs, bradycardia, asystole. 7 m Prone CPR then ROSC. PEA after 10 minutes ROSC, prone CPR x 4min then ROSC | Invasive BP with 40mmHg waveform correlated to compressions | Hemorrhage and venous air embolus in coronaries | Survived, ICU discharge day 7. Mild cognitive delay. |
| Loewenthal, 1993 | 53 | Female | Meningioma | Craniotomy | Asystole, prone CPR x 3min then ROSC | Palpable femoral pulse correlated to CPR | Venous air embolus | Survived |
| Mayorga-Buiza, 2018 | 10 | N/a | Posterior fossa tumor | Posterior fossa tumor excision | Pulseless SVT to VF. CPR x 8min then ROSC | Not discussed | Not discussed | Survived |
| Mishra, 2019 | 35 | Female | C2–C3 intradural extramedullary meningioma | Laminectomy and tumor excision | Rhythm not discussed. Prone CPR x1m, then flipped for supine CPR x2m then ROSC | Not discussed | Cardiac tamponade | Survived to discharge. |
| Sun, 1992 | 14 | Female | Open occipital fracture with left intracerebellar hematoma | Posterior fossa craniectomy | Bradycardia, PEA, idioventricular | Arterial line systolic pressure throughout: 100 to 160 mmHg . | Torn transverse sinus, blood loss, brain retraction progressing to severe bradycardia and hypotension | Survived, uneventful recovery |
| Sun, 1992 | 34 | Male | Compression fracture of third cervical vertebrae, incomplete severance of cervical spinal cord | Decompression laminectomy | PEA, VF | Arterial systolic pressure throughout: 120-200mmHg | Endotracheal tube occlusion | Survived, uneventful recovery |
| Sutherland, 1997 | 8 | Female | Limb girdle muscular dystrophy, progressive 55" thoraco-lumbar scoliosis | T1-sacrum posterior spinal fusion with sublaminar wiring | Asystole | CPR produced central venous pressure waveform but no arterial waveform | Air embolus | Deceased |
| Sutherland, 1997 | 12 | Female | Athetoid cerebral palsy, 90" progressive thoraco-lumbar scoliosis | T2-sacrum posterior spinal fusion with sublaminar wiring | Asystole | Not discussed | Air embolus | Deceased |
| Taylor, 2013 | 69 | Male | Metastatic melanoma and hypertrophic cardiomyopathy | posterior craniotomy | VT, progressed into VF | Pulsatile pressure tracing visible on the arterial line, and an ETCO<sub>2</sub> of 15 mmHg. | Unknown | Survived, full neurologic recovery |
| Tobias, 1994 | 12 | Male | Spastic quadriplegia, progressive scoliosis. | Spinal fusion | Bradycardia, asystole, idioventricular | Compressions pressures of 80-90 mm Hg from both arterial line and non-invasive BP cuff.. | >4L blood loss: abrupt decrease in MAP to 40 mmHg. | Survived, no change in neurologic status |
| Woo-Ming 1966 | 2 | Male | Whooping cough | n/a | No monitoring | Not discussed | Airway occlusion | Survived |
| Woo-Ming 1966 | 3 | Female | Stridor due to laryngeal diphtheria | Emergency tracheostomy | No monitoring | Not discussed | Laryngeal diphtheria | Survived, no deficits |
| Woo-Ming 1966 | 3 month | Male | Respiratory tract infection | n/a | No monitoring | Not discussed | Hypovolemia, airway occlusion | Survived |
| Woo-Ming 1966 | 7 weeks | Male | Staphylococcal pneumonia | n/a | No monitoring | Not discussed | Unknown | Survived |

Abbreviations:
PEA Pulseless electrical activity; VF ventricular fibrillation; SVT supraventricular tachycardia; M minute(s); ROSC return of spontaneous circulation; TTE transthoracic echocardiogram.

### Prone compression quality

Compression quality was assessed by invasive blood pressure monitoring, exhaled carbon dioxide and pulse palpation. A 2003 single-center case series reports of patients in intensive care with prolonged cardiac prone CPR was associated with improvements in both systolic (23±14 mmHg, p<0.05) and mean arterial blood pressure (14±11 mmHg, p<0.05) [37] In a fresh cadaver study, Wei et al (2006) found similarly greater systolic and diastolic blood pressures (SDP, DBP) in the prone (79 ± 20 /17 ± 10mmHg) versus supine (55 ± 20/13 ± 7mmHg) position p=0.028).[38] In five cases, prone compressions generated SBPs over 80mmHg.[18,22,32,35] In one case, SBP did not rise above 40mmHg in one case,[28] and in a cardiac arrest following air embolus, prone compressions produced a central venous but no arterial waveform.[33]

During craniotomy for acute cerebellar hemorrhage in an octogenarian, an EtCO_2_ of 33mmHg was achieved during prone compressions for an arrest linked to right-sided cardiac decompensation.[25] During a microscopic discectomy in a 43 year old, an EtCO_2_ level of 28mmHg was achieved during prone compressions following cardiac arrest of unknown cause.[23] Two additional cases reported EtCO_2_ of 15 and <10, [24,34] and AlHarbi reported lower EtCO2 in the prone position than supine.[17]

### Tidal volumes generated by prone compressions

One study examined the effect of prone compressions on tidal volumes.[38] In healthy volunteers, external back compressions generated mean tidal volumes of 399 +/-110mL in five male and five female volunteers.[38]

### Landmarking and hand position

In order to ascertain the best position for prone compressions, one study retrospectively reviewed chest computed tomography (CT) to correlate the surface landmarks overlying the left ventricle.[40] The largest left ventricle cross-section area was determined to be zero to two vertebral segments below the inferior angle of the scapula in at least 86% of patients in the prone position.[40]

### Survival

Twenty of the 25 cases (80%) were associated with post-resuscitation survival, although few cases included long term outcome.[17,18,20–24,26,28–32,34–36] The most commonly reported survival endpoint was alive at the end of surgery,[17–28,30–32,34] followed by survival to hospital discharge,[18,25,28,31] and seven cases reported full neurological recovery. [18,24,30,31,34–36]

## Discussion

Despite growing interest in prone positioning, most healthcare professionals are unfamiliar with prone CPR. Accordingly, any practice change would need widespread education and practice. This, in turn, should be justified by robust evidence. Our review highlights that there may be more supportive literature than many clinicians realize, but also many important gaps. The first recommendation for prone CPR was over three decades ago (1989),[41] meaning the concept is not new. What has changed is its importance: because more patients will be ‘proned’ due to covid-19, and because this practice may continue henceforth. In short, an increase in proning could mean more cardiac arrests in the prone position.

Presumably, the most pressing question is whether patients survive when resuscitated proned. After all, if universally unsuccessful then it should not be performed, or we need to accept/minimize the risks and delays associated with ‘supine-ing’ patients. The first concern is that the available guidance is predominantly from single cases or case series. Studies have also yet to widely encompass the sickest ICU patients. These ICU patients often have low post-arrest survival, because of multiorgan impairment and because the arrest commonly occurs despite aggressive preemption. This means that even before covid-19, ICU practitioners faced uncertainty regarding whether to perform default CPR. They now have added risk to themselves, and responsibility to keep their teams safe. Overall, practitioners must currently extrapolate from the operating room experience. Brown et al. reviewed 22 case reports of prone cardiac arrests, 10 of whom survived to discharge, [42] and Mazer et al. and Wei et al. found prone compressions generated a higher systolic and mean arterial pressure during circulatory arrest in ICU patients compared to standard CPR.[37][38]

In terms of guidelines, and despite the dearth of data, the UK Resuscitation Council does recommend starting chest compressions without changing the position for adult patients who arrest during neurosurgery.[43] They added that the efficacy of CPR should be judged using the end-tidal CO_2_ monitor, and the arterial pressure waveform. Prudently, they advise that patients be turned quickly supine if feedback from these monitors is judged insufficient. The 2010 American Heart Association guidelines for cardiopulmonary resuscitation and emergency cardiovascular care are more circumspect but still recommended: ‘when patients cannot be placed in the supine position, it may be reasonable for rescuers to provide CPR in the prone position, particularly in hospitalized patients with an advanced airway’.[11]

The next issue concerns optimum technique for prone chest compressions. The Resuscitation Council and American Heart Association make no specific recommendations. However, the first two cases of successful prone chest compressions (in neurosurgical patients) were published in 1992 by Sun et al, using what they called ‘reversed precordial compression’.[32] In everyday language, this means compressing over the mid-thoracic spine plus a hand under the lower sternum: for counter-pressure.[32] Similarly, Dequin et al. reported successful prone CPR in an ICU patient, in which they used two-handed mid-thoracic compression and a second person for sternal counter-pressure.[22] Gomes et al. reported successful prone CPR in a neurosurgical patient, using compression over the mid-thoracic level, but no sternal counter-pressure.[44] Kwon et al. reviewed 100 chest CTs to determine where to compress the largest left ventricular mass. Once again, it was mid-thoracic, below the inferior angle of the scapulae.[40]

In 2020, guidelines were expedited by the UK’s Faculty of Intensive Care Medicine and Intensive Care Society, spurred by the covid-19 pandemic.[10] These organizations jointly recommend a two-handed technique over the mid-thoracic spine, and between the two scapulae. The use of a second person to apply counter-pressure is optional. Given the added logistics, the contamination risk, and the overall recommendation to limit resuscitator in the room, we would recommend against this additional step.

Clearly, our scoping review has limitations which limit its generalizability. These include a general lack of studies, a focus on operating theatre patients and a preponderance of witnessed arrests. The evidence is mostly from cases, and is therefore of comparatively low quality and predates the 2015 resuscitation guidelines. We also had to rely upon Google Translate for one French, one German and one Korean case report. Regardless, rather than prone resuscitation being as an area of niche interest, we believe our modest scoping review has highlighted an area of increasingly clinical relevance and research priority.

### Conclusion

This scoping review demonstrated that the majority of published literature on prone resuscitation are neurosurgical cases with positive outcomes. Although this scoping review has not identified sufficient evidence to prompt a systematic reviews or reconsideration of current guidelines, it pinpoints gaps in the research evidence related to prone resuscitation, namely a lack of moderate to high-level evidence and paucity of studies of out-of-hospital cardiac arrest. We hope this review can help inform the care of prone patients during the covid-19 pandemic should they require resuscitation.

## Data Availability

All relevant data is included in the article and appendices.

